# Functional alteration of innate T cells in critically ill Covid-19 patients

**DOI:** 10.1101/2020.05.03.20089300

**Authors:** Youenn Jouan, Antoine Guillon, Loïc Gonzalez, Yonatan Perez, Stephan Ehrmann, Marion Ferreira, Thomas Daix, Robin Jeannet, Bruno François, Pierre-François Dequin, Mustapha Si-Tahar, Thomas Baranek, Christophe Paget

## Abstract

Covid-19 can induce lung infection ranging from mild pneumonia to life-threatening acute respiratory distress syndrome (ARDS). Dysregulated host immune response in the lung is a key feature in ARDS pathophysiology. However, cellular actors in Covid-19-driven ARDS are poorly understood. Here, we dynamically analyzed the biology of innate T cells, a heterogeneous class (MAIT, γδT and iNKT cells) of T lymphocytes, presenting potent anti-infective and regulatory functions. Patients presented a compartmentalized lung inflammation paralleled with a limited systemic inflammation. Circulating innate T cells of critically ill Covid-19 patients presented a profound and persistent phenotypic and functional alteration. Highly activated innate T cells were detected in airways of patients suggesting a recruitment to the inflamed site and a potential contribution in the regulation of the local inflammation. Finally, the expression of the CD69 activation marker on blood iNKT and MAIT cells at inclusion was predictive of disease severity. Thus, patients present an altered innate T cell biology that may account for the dysregulated immune response observed in Covid-19-related acute respiratory distress syndrome.

## Introduction

In December 2019 were first reported in Wuhan, China, pneumonia cases due to a coronavirus, the Severe Acute Respiratory Syndrome coronavirus 2 (SARS-CoV-2), a novel strain related to SARS-CoV and MERS-CoV, responsible for recent outbreaks. Disease presentation related with SARS-CoV-2 (Coronavirus disease, Covid-19), can vary from mild disease to life-threatening acute respiratory distress syndrome (ARDS). ARDS is caused by a sustained and dysregulated immune response triggered in the lung after initial insult, resulting in alteration of alveolar-capillary membrane permeability and perturbed tissue repair^1^. This pathological process leads to interstitial and alveolar oedema that profoundly impairs gas exchange. However, the cellular and molecular factors that are responsible for this aberrant and persistent inflammatory response are poorly understood^2^. In SARS-CoV infection, delayed type I IFN response together with high viral loads were associated with defective adaptive response and exaggerated tissue damage^3^. During severe SARS-CoV-2 infection, elevated pro-inflammatory cytokines levels (*e.g*. IL-6 and TNF-α) were associated with more severe cases, supporting an inflammatory hypothesis^4–6^. In addition, T cell lymphopenia has been correlated with disease severity suggesting a role for these cells in the pathophysiology of severe Covid-19^5,6^. Besides classical adaptative CD4^+^ and CD8^+^ T cells, the T cell compartment comprises several lineages of cells endowed with both innate and adaptive properties that are referred to as unconventional or innate T cells (iT cells)^7^. This heterogeneous class of T cells comprises three main lineages including Mucosal-Associated Invariant T (MAIT), γδT and invariant Natural Killer T (iNKT) cells. They are restricted to quasi-monomorphic non-classical major histocompatibility complex and have emerged as key players in mucosal immunity and inflammatory response^8–11^. Given their versatile functions, iT cells emerge as interesting targets in the context of Covid-19-driven ARDS. First, iT cells mainly populate mucosal tissues including the lung and have the ability to promptly produce substantial amounts of inflammatory cytokines such as IFN-γ and IL-17A, two key cytokines in anti-infective response at barrier sites. Moreover, iT cells can fine-tune the intensity and flavour of the host immune response shaping the magnitude of the adaptive response. They can also participate in the process of the resolution of inflammation including tissue repair and regeneration^12–14^, a critical step that is highly altered during ARDS. Despite these pivotal functions, the putative contribution of iT cells in the pathophysiological process of ARDS has never been explored.

Here, we dynamically assessed the relative frequencies and functions of iT cells in biological fluids of thirty patients with severe Covid-19 admitted to intensive care unit (ICU). Our analysis indicated that iT cells from critically ill Covid-19 patients displayed a phenotype of activated cells associated with changes in their cytokine profile. Importantly, activated iT cells populated the airways of patients presenting a strong local inflammation. In addition, the activation status of blood iT cells on admission was predictive of the level of hypoxia during the course of infection. Thus, our study indicates that an alteration in iT cell biology may account for the sustained host immune response dysregulation observed in Covid-19-driven ARDS.

## Results and Discussion

### Lymphopenia and compartmentalized lung inflammation characterized critically ill Covid-19 patients

Thirty patients admitted in ICU for severe Covid-19 were included. Baseline characteristics of the patients are presented in **Table 1**. Median duration of symptoms before admission in ICU was 10 days (8; 14), and ultimately, 24 patients (80 %) required invasive mechanical ventilation (20 at admission). Among these mechanically ventilated patients, all presented an ARDS, 21 (70 %) received neuromuscular blockade, 18 (60 %) were placed on prone position and one patient required extra-corporeal membrane oxygenation. On CT scan, we observed typical bilateral diffuse ground-glass lesions in severe Covid-19 patients (**Supplementary Figure 1A**). Upon enrolment, most of Covid-19 patients (22/30) presented mild to severe lymphopenia (0.75 × 10^9^/L ± 0.06) (**Figure 1A**) accompanied by a higher neutrophil-to-lymphocyte ratio as compared to age- and sex-matched controls (**Figure 1B**). As reported^15^, we observed a positive correlation between the degree of lymphopenia and the Sequential Organ Failure Assessment (SOFA) score (**Figure 1C**). Similarly, the lymphopenia was more pronounced in patients under invasive mechanical ventilation as compared to patients who did not require invasive mechanical ventilation (**Figure 1D**). Among the 30 critically ill Covid-19 patients, one died at day 2 after inclusion and 14 (46.7%) were still in ICU at day 15, including 9 still under invasive mechanical ventilation. The remaining 15 patients improved and were discharged from ICU to other wards.

**Table 1:** Clinical and laboratory information of the study patients.

|  |  |
| --- | --- |
| Age (years), median (IQR) | 64 (57; 67) |
| Male / Female ratio | 2/1 |
| Body Mass Index (kg/m<sup>2</sup>) | 31 (28; 32) |
| **Comorbidities** |  |
| Hypertension, n (%) | 12 (46.7) |
| Type 2 diabetes | 9 (30) |
| Active smoking | 0 |
| Chronic respiratory failure | 0 |
| Chronic renal failure | 1 (3.3) |
| Cardiovascular disease | 3 (10) |
| Immunosuppression | 1(3.3) |
| SAPS2 | 32 (22; 36) |
| SOFA | 4 (2; 6) |
| Invasive mechanical ventilation on admission | 20 (66.7) |
| **Laboratory data** |  |
| Lymphocytes (x10<sup>9</sup>/L), median (IQR) | 0.715 (0.510; 0.998) |
| Neutrophils (x10<sup>9</sup>/L), median (IQR) | 6.25 (4.3; 7.52) |
| CRP (mg/L), median (IQR) | 187.7 (83.8; 258) |

**Figure 1:**
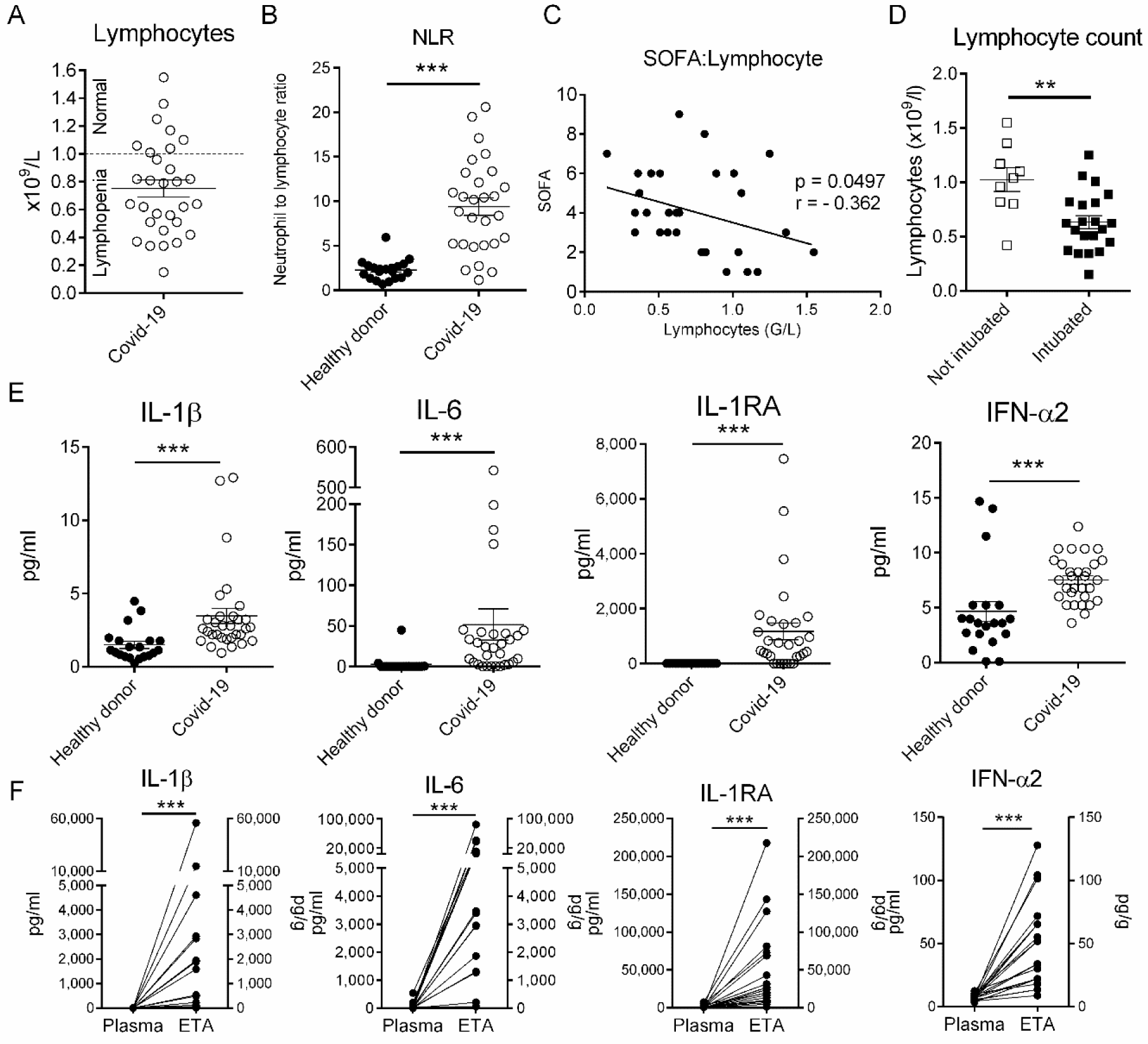
Inflammatory status of critically ill patients with Covid-19. **A**, Lymphocyte count in whole blood from critically ill Covid-19 patients (n = 30) was analyzed at day 1 post admission. Individuals and means ± SEM are depicted. **B**, Neutrophil-to-lymphocyte ratio in the whole blood of healthy donors (n = 20) and patients (n = 30) was determined by flow cytometry. Individuals and means ± SEM are shown. **C**, Spearman’s rank correlation of SOFA and lymphocyte counts in Coiv-19 patients. **D**, Lymphocyte counts at day 1 in patients with (n = 21) or without (n = 9) mechanical ventilation. Individuals and means ± SEM are depicted. **E**, Levels of IL-1β, IL-6, IL-1RA and IFN-α2 in the plasma of healthy donors (n = 10) and severe Covid-19 patients (n = 30). Individuals and means ± SEM are shown. **F**, Levels of IL-1β, IL-6, IL-1RA and IFN-α2 in the plasma and ETA supernatants of matched patients. Paired individual values are shown. **, p<0.01; ***, p<0.001.

Although the circulating levels of the inflammatory mediators IL-1β and IL-6 were significantly higher in Covid-19 patients as compared to age- and sex-matched controls (**Figure 1E**), the detected amounts were relatively low, as previously reported^6^. Of note, the levels of plasma IL-1RA were high suggesting an active anti-inflammatory process (**Supplementary Figure 1B**). As judged by IFN-α2 levels, the type I IFN response, a critical component of the anti-viral response^16^ was also low in the blood compartment of patients as compared to controls (**Figure 1E**). To monitor the local inflammation, we analyzed the same mediators in the supernatants of endotracheal aspirates (ETA) of matched Covid-19 patients (**Figure 1E**). Strikingly, the amounts of IL-6, IL-1β and to a lesser extent IFN-α2 were sky high in the airways suggesting an intense local inflammation upon ICU admission (**Figure 1F**). Collectively, our data indicate a compartmentalized inflammation in patients with severe Covid-19.

### Blood innate T cell decrease in Covid-19 patients is paralleled with their presence in the airways

While a T cell lymphopenia has been described in severe Covid-19 patients^6^, the innate T cell compartment has not been investigated yet. Detailed analysis of circulating iT cells within total T lymphocytes indicated a profound decrease in MAIT (~ 6-fold) and iNKT (~ 7-fold) relative proportions in critically ill Covid-19 patients (**Figure 2A**) despite some comorbidities such as obesity and/or diabetes can also partly contribute to this observation^17,18^. In the meantime, the relative proportion of γδT cells remained unchanged (**Figure 2A**). However, by focusing on subsets based on TCR repertoire, we observed that the frequency of Vδ2^+^ was slightly decreased while the Vδ1δ2^−^ cells were increased (**Figure 2A**). Of note, no significant differences could be observed for the Vδ1^+^ subset (**Figure 2A**). This may indicate a particular contribution of this γδT subset - likely Vδ3^+^ - during Covid-19 as suggested in other viral infections^18^.

**Figure 2:**
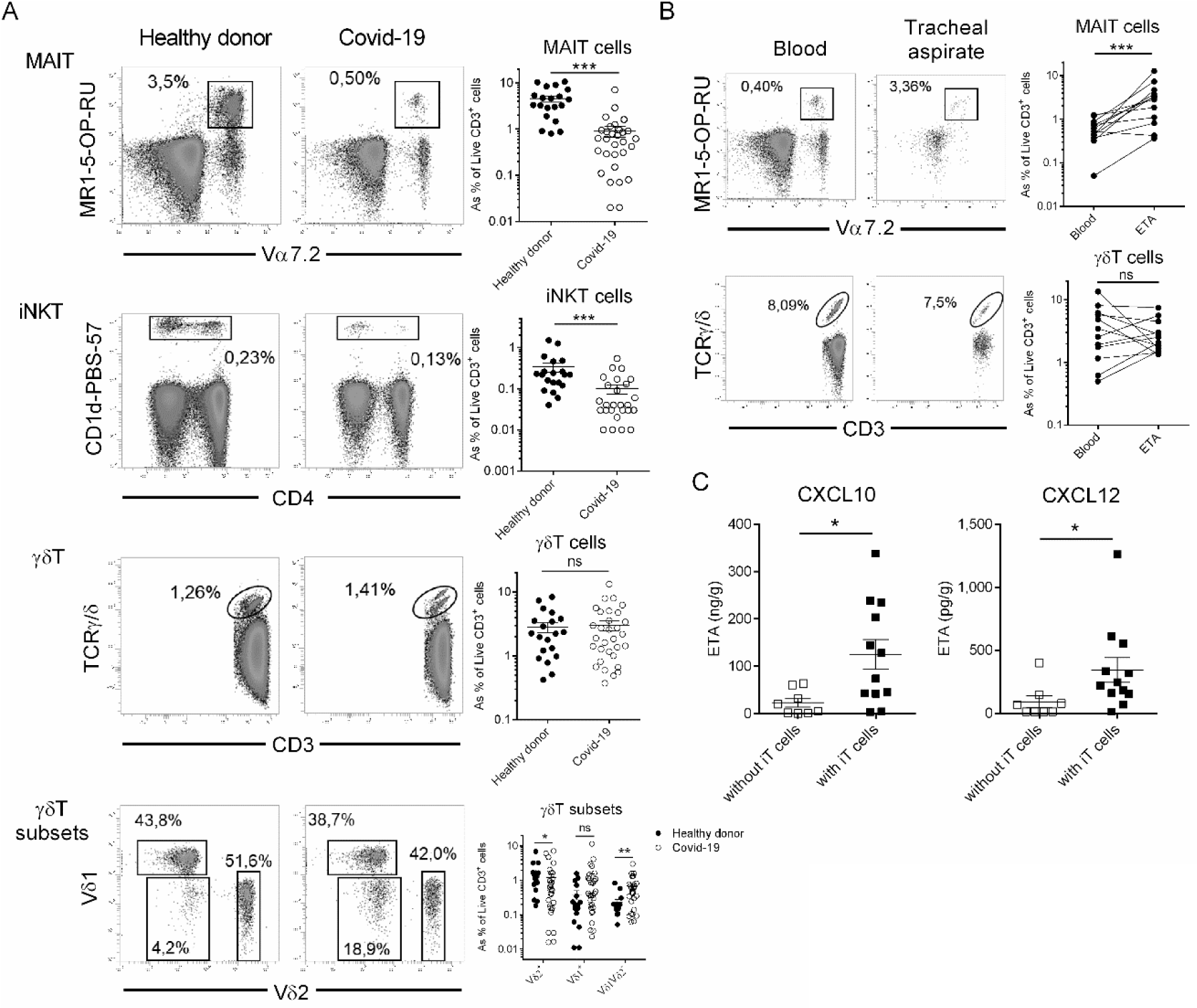
Relative proportion of innate T cells in PBMC and ETA of Covid-19 patients. **A**, Flow cytometry analyses of innate T cells in the blood of healthy donors (n = 20) and severe Covid-19 patients (n = 30). Representative dot plots of MAIT, iNKT and γδT cells from healthy donors and Covid-19 patients as percentage of CD3^+^ live cells are shown in the left panel. Proportion of γδT subsets is shown as percentage of total γδT cells. Individuals and means ± SEM are shown in the right panel. Of note, iNKT cells could not be detected in 4 patients. **B**, Comparative analysis of MAIT and γδT cell subsets in blood and ETA of 12 Covid-19 patients with analyzable lymphocyte compartment in ETA. Representative dot plots are shown in the left panel. Individuals and means ± SEM are shown in the right panel. **C**, Levels of CXCL10 and CXCL12 in ETA supernatants according to the presence (n = 12) or not (n = 8) of iT cells. Individuals and means ± SEM are shown. ns, not significant; *, p<0.05; ***, p<0.001.

The decrease in circulating MAIT and iNKT cells may have multiple causes. First, we interrogated whether this could be a consequence of activation-induced TCR internalization. However, mean fluorescence intensity for TCR expression indicated no modulation for the iNKT TCR between patients and controls (**Supplemental Figure 2A**) while an increase intensity was observed for the MAIT TCR of patients. In addition, the levels of intracellular TCR Vα7.2 expression in T cells of patients did not indicate any signs of MAIT TCR internalization (**Supplemental Figure 2B**).

Another explanation could be their migration into the airways of patients. Thus, we monitored the putative presence of iT cells in ETA of Covid-19 patients under mechanical ventilation (**Table 1**). We were able to recover cells - containing more than 1% of lymphocytes - in 12 ETA samples out of 21, enabling a further analysis of iT cells. MAIT and γδT cells were detected in airways of all patients with an analysable lymphocyte compartment (**Figure 2B**). Of note, airway iNKT cells were virtually undetectable in all samples. Interestingly, the frequency of MAIT cells but not of γδT cells was higher in the airways as compared to blood of matched-patients (**Figure 2B**) suggesting that the presence of MAIT cells in the airways might be dependent on an active recruitment mechanism rather than blood vessel leakage due to the alveolar-capillary barrier disruption. Furthermore, the presence of iT cells in ETA was associated with a higher level of the chemoattractants for T cells CXCL10 and CXCL12^19^ (**Figure 2C**). Of note, one could argue that other causes such as increased cell death might account in this phenomenon^20^. Altogether, these data indicate a decrease in circulating MAIT and iNKT cells that may be a consequence of their recruitment into the airways.

### Innate T cells displayed an altered functional profile in Covid-19 patients

Phenotypic analysis of iT cell in blood of Covid-19 patients showed an increased expression of the activation marker CD69 in all subsets (**Figure 3A**). This phenotype was associated with a higher level of IL-18 in the plasma of patients (**Supplemental Figure 3**), a cytokine associated to iT cell activation during viral infections^21–23^. In parallel, we observed an increased level of blood PD-1-expressing iT cells in patients suggesting a certain level of exhaustion (**Figure 3A**). Of importance, the levels of CD69 and PD-1-expressing MAIT and γδT cells were significantly higher in ETA compared to blood in matched patients (**Figure 3B**), which further support the possible migration of activated blood iT cells into the lungs.

**Figure 3:**
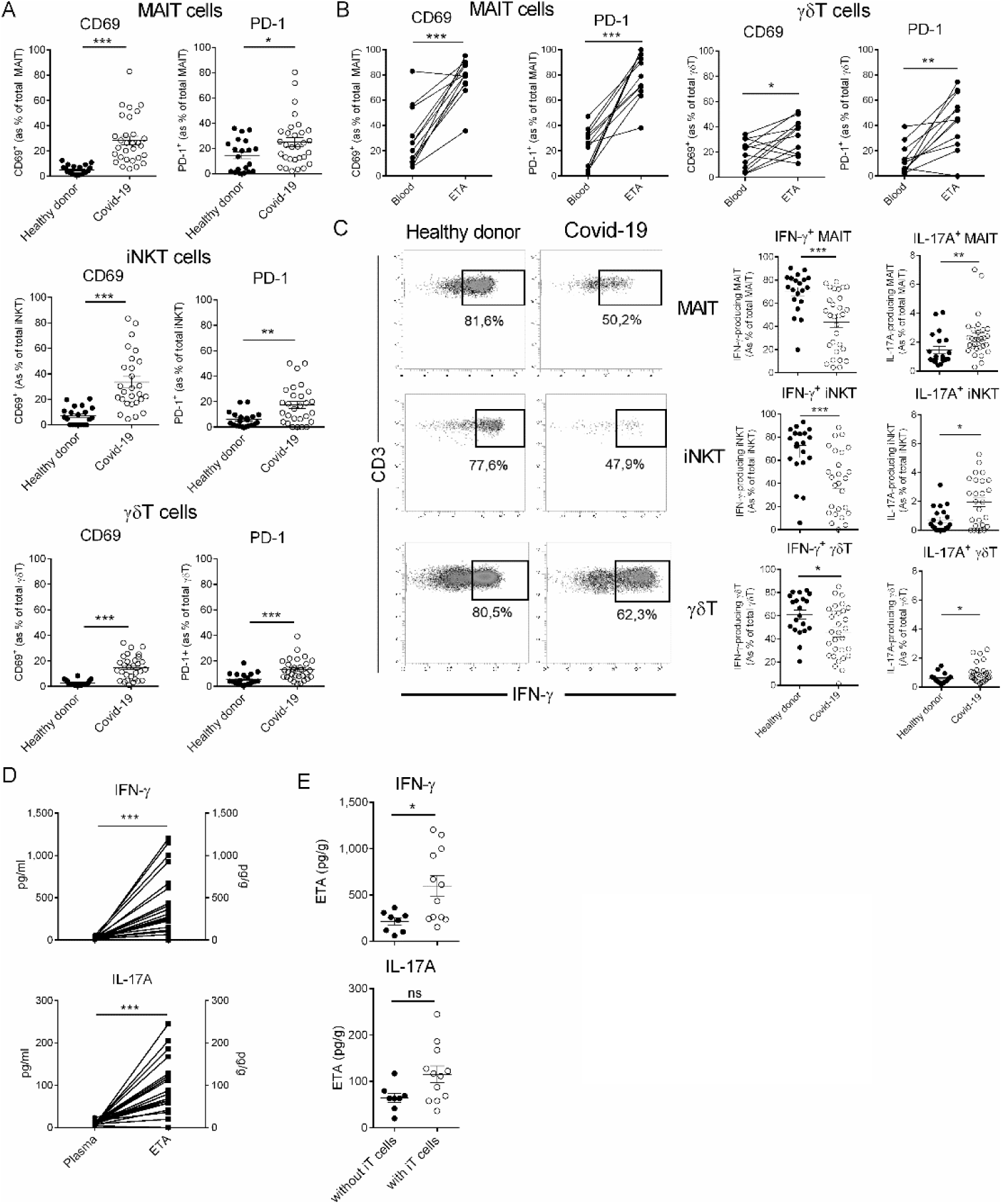
Functional analysis of innate T cells during severe Covid-19. **A**, Flow cytometry analyses of CD69 and PD-1 expression on MAIT, γδT and iNKT cells in the blood of healthy donors (n = 20) and severe Covid-19 patients (n = 30). Individuals and means ± SEM are shown. **B**, Relative proportions of CD69^+^ and PD-1^+^ iT cells in the blood and ETA of matched patients. Paired individual values are shown. **C**, Intracellular staining for IFN-γ and IL-17A of PMA/ionomycin-activated PBMC. Representative dot plots for iT cells are depicted in the left panel. Individuals and means ± SEM are shown on the right panel. **D**, Levels of IFN-γ and IL-17A in the plasma and ETA supernatants of matched patients. Paired individual values are shown. **E**, Levels of IFN-γ and IL-17A in the ETA supernatants according to the presence (n = 12) or not (n = 8) of iT cells. ns, not significant; *, p<0.05; **, p<0.01; ***, p<0.001.

In addition, cytokine production by fresh blood iT cells was analyzed after short-term PMA/ionomycin stimulation. Circulating MAIT and iNKT, and to a lesser extent γδT cells from Covid-19 patients produced less IFN-γ as compared to cells from healthy donors (**Figure 3C**). In the meantime, this was paralleled with an increased ability of iT cells from patients to produce IL-17A (**Figure 3C**) although the levels detected were relatively low. This could be partially explained by the migration of IFN-γ-producing iT cells to the lung tissue. In line with the acute inflammation in the lung of Covid-19 patients^24^, the levels of IFN-γ and IL-17A were more elevated in the supernatants of ETA compared to plasma of matched patients (**Figure 3D**). Interestingly, they were present at higher concentration in iT cell-containing ETA samples (**Figure 3E**) suggesting that iT cells may contribute to the production of IFN-γ and IL-17A in airways of critically ill Covid-19 patients. Collectively, our data indicate a functional alteration of iT cells in Covid-19 patients.

### Alteration in innate T cell biology is persistent and correlates with disease severity

To gain insight into their temporal changes during Covid-19, the frequency and phenotype of blood iT cells was monitored in patients during their stay in the ICU. We observed a decrease in the frequency of MAIT cells at day 7 and then it was maintained to day 14 (**Figure 4A**). The proportion of CD69- and PD-1-expressing MAIT cells followed a similar trend (**Figure 4A**). The relative proportion of circulating iNKT cells was stable in critically ill Covid-19 patients during their stay in ICU regardless of their clinical course (**Figure 4B**). The level of CD69-expressing iNKT cells was reduced at day 7 and 14 while no significant changes could be observed for PD-1 expression on iNKT cells while (**Figure 4B**). Similar to MAIT cells, the relative proportion of γδT cells in was reduced at day 7 and a further decrease was noticed at day 14 (**Figure 4C**). Regarding their phenotype, the levels of circulating PD-1-expressing γδT cells was stable in patients while the relative proportion of their CD69-expressing counterparts was decreased at day 7 and 14 (**Figure 4C**).

**Figure 4:**
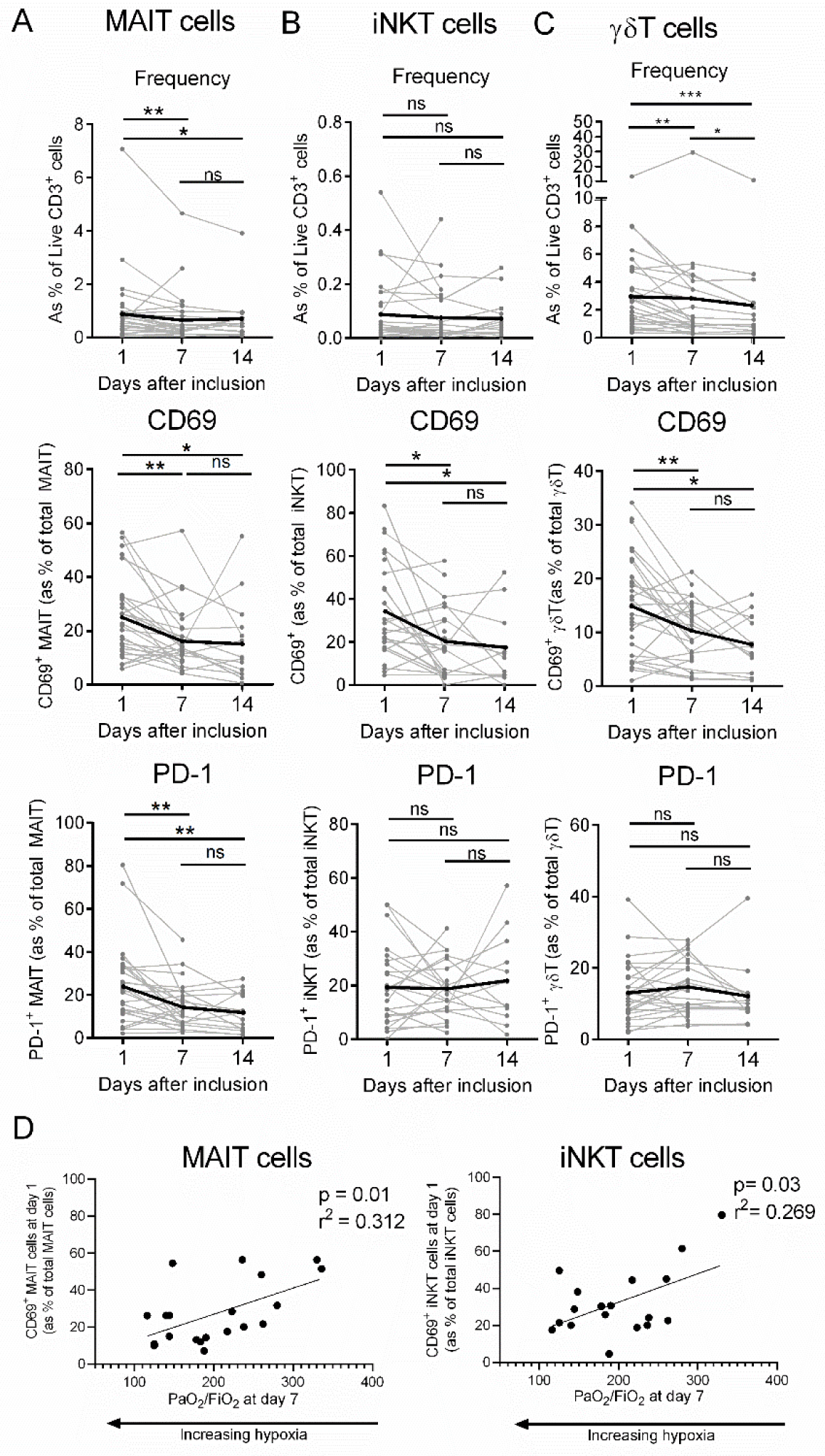
Kinetic analysis of the frequency and phenotype of innate T cells in severe Covid-19 patients. **A-C**, Flow cytometry analyses of relative proportion and CD69 and PD-1 expression on MAIT (A), γδT (B) and iNKT (C) cells in the blood of critically ill Covid-19 patients at days 1 (n = 30), 7 (n = 27) and 14 (n = 14). Kinetics plots showing mean value for each patient (each grey line corresponds to one patient). Median values for each parameters were plotted in black. **D**, Spearman’s rank correlation of CD69 expression on blood iT cells and hypoxia levels in Covid-19 patients. ns, not significant; *, p<0.05; **, p<0.01; ***, p<0.001.

Finally, we interrogated whether alterations in iT cell biology could be predictive of the clinical course of critically ill Covid-19 patients especially regarding the level of hypoxia, measured by the PaO_2_/FiO_2_ ratio, a routine clinical variable for ARDS management^1^. Thus, we compared multiple iT cell parameters on admission to the level of hypoxia at day 7. Although no correlation could be observed for γδT cells, we observed that CD69 expression on MAIT and iNKT cells was positively correlated to the PaO_2_/FiO_2_ ratio indicating a reduced level of hypoxia (**Figure 4D**). Notably, no additional correlation could be observed for MAIT and iNKT cells based on the other iT cell-related parameters including PD-1 expression and cytokine production.

Altogether, these data indicate a persistent alteration in iT cell activation in Covid-19 patients and the levels of activation on MAIT and iNKT cells upon admission can be predictive of disease severity.

## Methods

### Clinical study design, patient population and approval

Patients (>18 years old) admitted in ICU with positive SARS-CoV-2 RT-PCR testing were prospectively included in this study, from March 18^th^ 2020 to April 17^th^ 2020. The study was conducted in one ICU from an academic hospital (Tours, France). All patients or their next of kin gave consent for participation in the study. This work was part of an ongoing study exploring immune response during community-acquired pneumonia (ClinicalTrial.gov identifier: NCT03379207). The study was approved by the ethic committee “Comité de Protection de Personnes Ile-de-France 8” under the agreement number 2017-A01841-52, in accordance with the national laws. Blood samples from healthy volunteers (age- and sex-matched) were obtained from the “Etablissement Français du Sang”.

### Reagents and antibodies

Staining was performed using antibodies (**Table S1**) from Biolegend (San Diego, CA, USA) and Miltenyi Biotec (Bergish Gladbach, Germany). PBS-57 glycolipid-loaded and control CD1d tetramers (BV421-conjugated) as well as 5-OP-RU-loaded and control MR1 tetramers (BV421-conjugated) were obtained from the National Institute of Allergy and Infectious Diseases Tetramer Facility (Emory University, Atlanta, GA). Dead cells were stained with LIVE/DEAD® Fixable Aqua Dead Cell Stain kit (ThermoFisher Scientific, Illkirch, France).

### Human cell isolation

#### PBMC

Peripheral Blood Mononuclear Cells (PBMC) were enriched by density gradient centrifugations using Histopaque-1077 solution (Sigma-Aldrich) according to the manufacturer’s instructions. Red blood cells were removed using a red blood cell lysis buffer (Sigma-Aldrich).

#### Endotracheal aspirates (ETA)

ETAs were collected from Covid-19 patients who were under invasive mechanical ventilation. Then, ETAs were weighted and incubated in PBS (5 ml/g) with 1 mM Dithiothreitol for 30 min at 4°C under gentle agitation. After centrifugation, supernatants were collected and cell pellets were filtered through a 100 μm cell strainer. Red blood cells were removed using a red blood cell lysis buffer and then cells from ETA were passed through a 40 μm cell strainer prior staining for flow cytometry.

### Cytokine measurement

Cytokines were measured in sera and supernatants of ETA using the Bio-Plex Pro Human cytokines screening panel (Bio-Rad, Marnes-la-Coquette, France) in a multiplex fluorescent bead assay (Luminex), according to the manufacturer’s instructions.

### Flow cytometry

Cells were stained with antibodies to surface epitopes and viability dye (LIVE/DEAD Fixable Aqua Dead Cell Stain). For cytokine profile analysis, cells were stimulated for 4 h in RPMI1640 complete medium containing PMA (100 ng/ml) and ionomycin (1 μg/ml) in presence of protein transport inhibitor cocktail (eBioscience) added 1 hour after stimulation. For intracellular staining, cells were fixed and permeabilized using the Fixation/Permeabilization Solution Kit (BD Biosciences). Cells were stained with APC-conjugated mAb against IL-17A and PE-conjugated mAb against IFN-γ. Events were acquired on a MACS Quant (Miltenyi Biotec) cytometer. Analyses were performed by using the VenturiOne software (Applied Cytometry; Sheffield, UK).

### Statistical analysis

All statistical analysis was performed by using GraphPad Prism software. The statistical significance was evaluated by using non-parametric unpaired Mann-Whitney *U* tests in order to compare the means of biological replicates in each experimental group. In some cases, the non-parametric Wilcoxon matched-pairs signed rank test was used. Results with a P value of <0.05 were considered significant. ns: not significant; *p < 0.05; **p < 0.01; ***p < 0.001.). Correlation calculation between two parameters has been performed using the Spearman’s rho test.

## Data Availability

All data referred to in the manuscript are available upon request to

## Author contributions

Y.J., A.G., S.E., P-F.D., M.S., T.B. and C.P. designed the research; L.G., Y.P., M.F., T.D., R.J., T.B. and C.P collected the data; Y.J., L.G., T.B. and C.P. analysed the data; Y.J. and C.P. wrote the manuscript with the input of all authors.

## Acknowledgments

This work was supported by the Agence Nationale de la Recherche “JCJC program” (ANR-19-CE15-0032-01) and by the Fondation du Souffle, with the “Fond de recherche en Santé Respiratoire”. M.S. and C.P. are supported by Inserm. Y.J., A.G., Y.P., S.E. and P-F.D. were supported by the CHU of Tours. M.F., T.B. and L.G. are supported by the University of Tours. We thank the NIH tetramer core facility (Emory University) for providing CD1d tetramers. We acknowledge all healthcare co-workers involved in the ICU department at the Bretonneau Hospital, especially Aurélie Aubrey, Delphine Chartier, Véronique Siméon and Julien Bontemps for their excellent management of patient samples and clinical data. Annick Legras, Denis Garot, Emmanuelle Mercier, Charlotte Salmon-Gandonnière, Laetitia Bodet-Contentin, Marlène Morisseau, Stephan Mankikian and Walid Darwiche are acknowledged for patient inclusions. We thank all the patients and their families for their trust and confidence in our work.

---

**Supplemental Figure 1:**
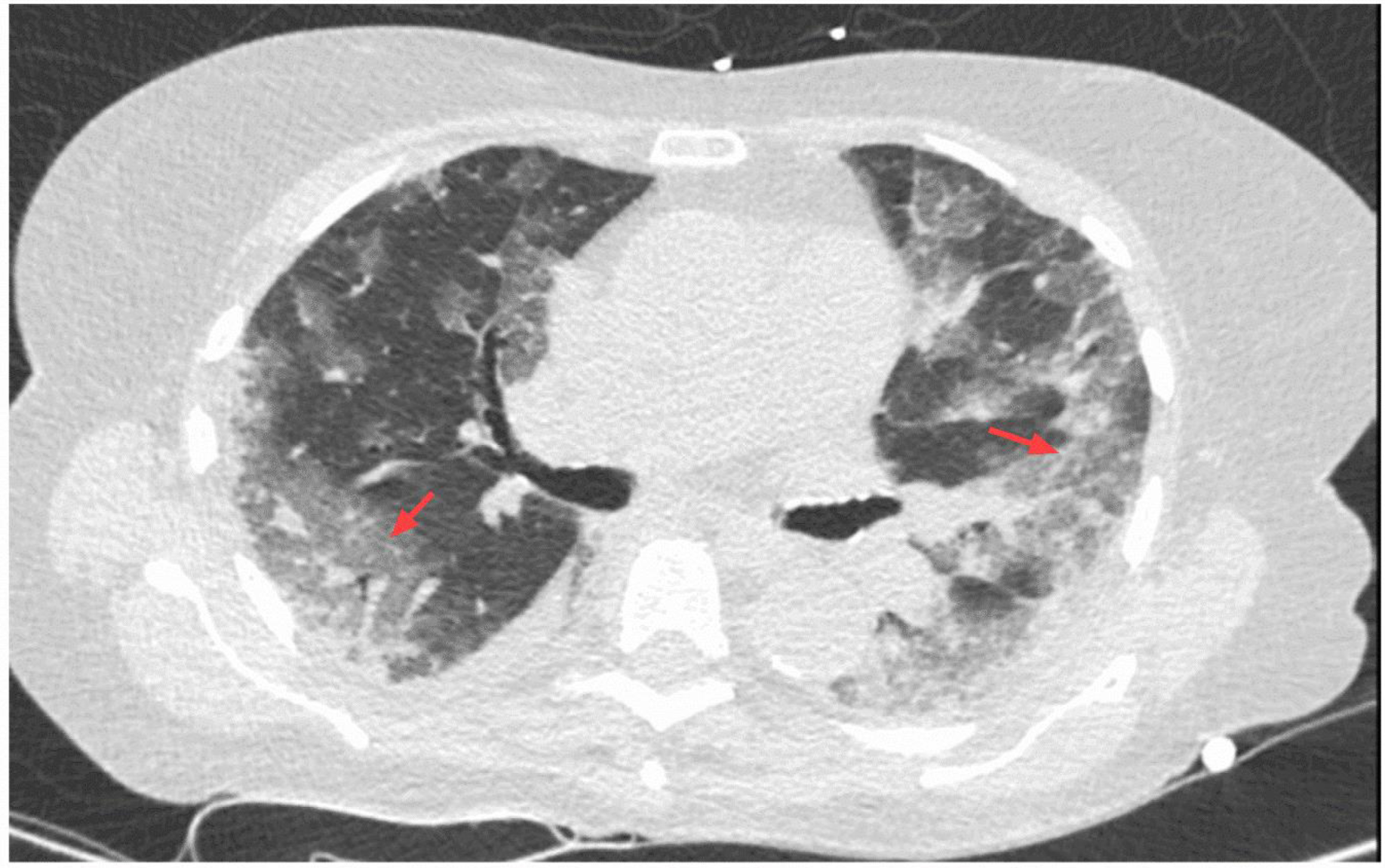
Chest computed tomography scan of Covid-19 patients. Representative thoracic computed tomography scanning of healthy volunteer and critically ill Covid-19 patient.

**Supplemental Figure 2:**
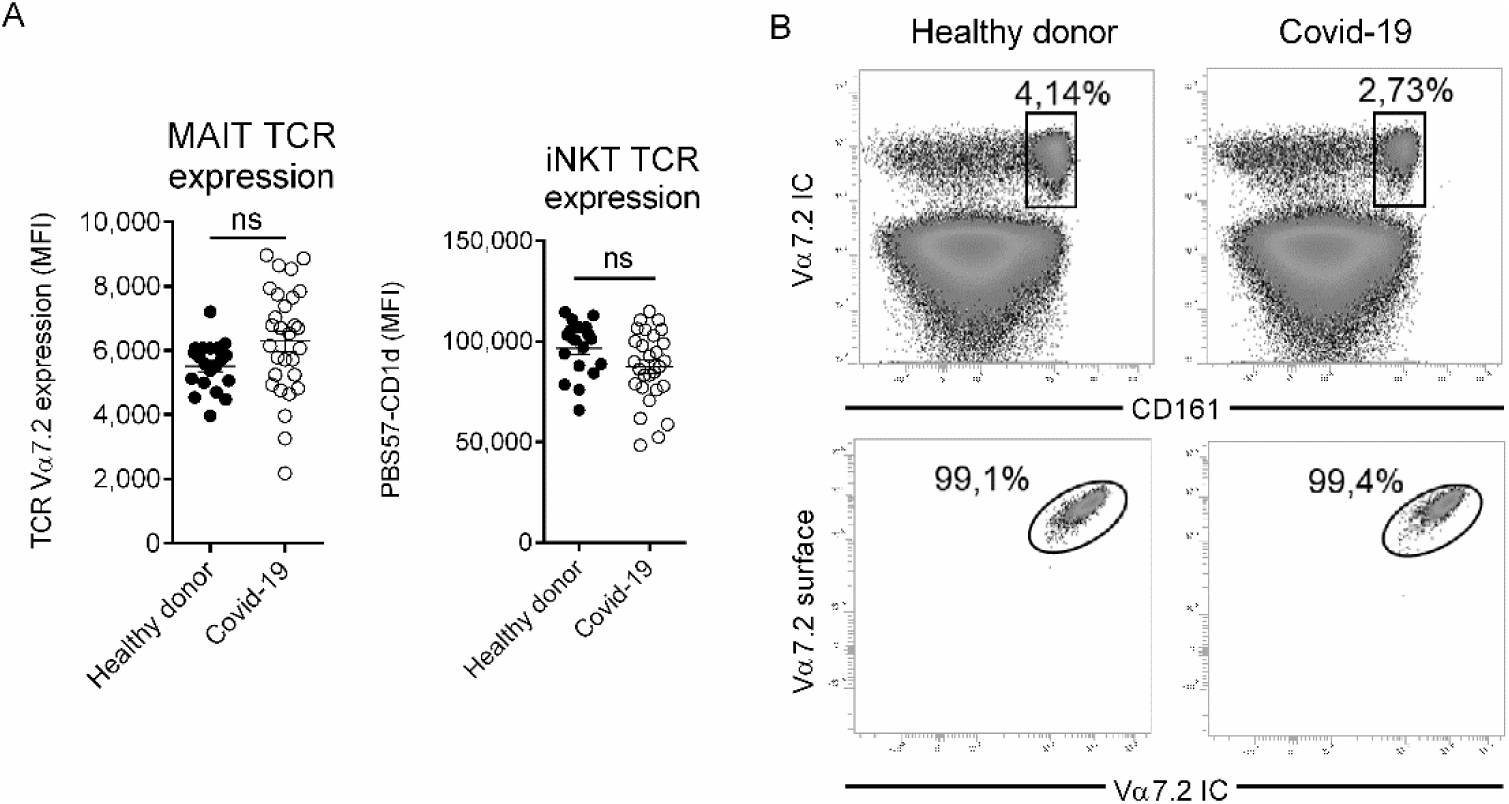
Evaluation of TCR internalization in MAIT and iNKT cells. **A**, Mean intensity fluorescence of TCR expression on MAIT and iNKT cells from control (n = 20) or Covid-19 patients (n = 30) based on TCR Vα7.2 and PBS57-CD1d tetramer staining respectively. Individuals and means ± SEM are shown. **B**, Flow cytometry analysis of TCR Vα7.2 expression using surface *vs* intracellular staining. Representative dot plots of 4 control and 4 Covid-19 patients are shown. ns, not significant; ***, p<0.001.

**Supplemental Figure 3:**
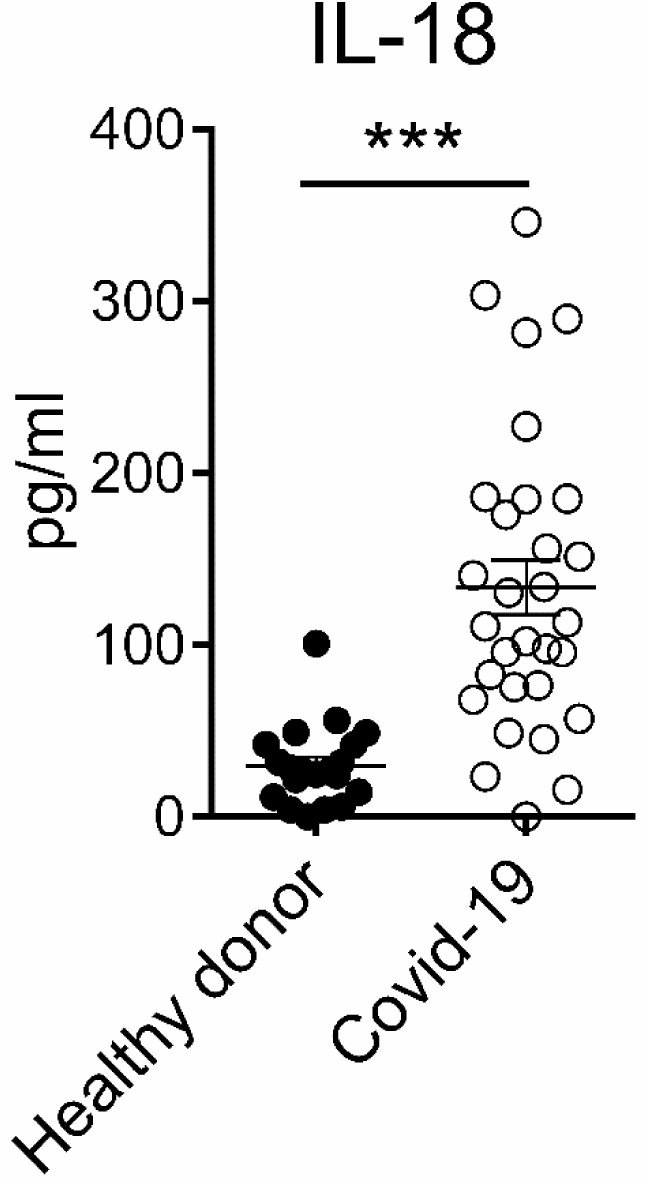
Plasma levels of IL-18 in critically ill Covid-19 patients. Levels of IL-18 in the plasma of healthy donors (n = 10) and severe Covid-19 patients (n = 30). Individuals and means ± SEM are shown. ***, p<0.001.

**Supplemental Table 1:** List of anti-human monoclonal antibodies

| **Epitopes** | **Clone** | **Fluorophore(s)** | **Manufacturers** |
| --- | --- | --- | --- |
| **CD3** | OKT3 | APC | BioLegend |
| **IL-17A** | BL168 | APC | BioLegend |
| **CD45** | 2D1 | APC/Cy7 | BioLegend |
| **CD4** | OKT4 | APC/Cy7 | BioLegend |
| **TCR Vδ2** | B6 | FITC | BioLegend |
| **TCR Vα7.2** | 3C10 | FITC | BioLegend |
| **CD3** | HIT3a | FITC | BioLegend |
| **IFN-γ** | 4S.B3 | PE | BioLegend |
| **CD69** | FN50 | PE | BioLegend |
| **PD-1** | A17188B | PE-Cy7 | BioLegend |
| **CD161** | HP-3G10 | PerCP | BioLegend |
| **TCR $\gamma/\delta$** | B1 | PerCP-Cy5.5 | BioLegend |
| **TCR Vδ1** | REA173 | VioBlue | Miltenyi Biotec |

